# Big tau and brain-derived tau reveal peripheral and central nervous system involvement in neuropathies

**DOI:** 10.64898/2026.08.27.26361202

**Authors:** Lorena Martín-Aguilar, Fernando González-Ortiz, Henrik Zetterberg, Thomas K. Karikari, Marc Suárez-Calvet, Carlos Casasnovas, Gerardo Gutiérrez-Gutiérrez, Maria José Sedano-Tous, Julio Pardo-Fernandez, Celedonio Márquez-Infante, Iñigo Rojas-Marcos, Ivonne Jericó-Pascual, Eugenia Martínez-Hernández, Germán Morís de la Tassa, Cristina Domínguez-González, Teresa Sevilla, Ana Lara Pelayo, Ricard Rojas-García, Roger Collet-Vidiella, Helena Codes-Méndez, Marta Caballero-Ávila, Clara Tejada-Illa, Cinta Lleixà, Gemma Riesco-Navarro, Nerea Blanco-Sanroman, Tania Mederer-Fernández, Lara Panicot-Buj, Elba Pascual-Goñi, Angela Vidal-Jordana, Kaj Blennow, Hlin Kvartsberg, Luis Querol

## Abstract

**INTRODUCTION:** Biomarkers for monitoring disease activity and treatment response in peripheral neuropathies remain limited. Big tau, a high-molecular-weight isoform of tau, is predominantly expressed in the peripheral nervous system (PNS). We investigated serum levels of big tau, brain-derived tau (BD-tau), and neurofilament light chain (NfL) in peripheral neuropathies, multiple sclerosis (MS), Alzheimer’s disease (AD), and healthy controls (HC).

**METHODS:** Ultra-sensitive blood-based assays run on an HD-X Single Molecule Array analyser (Quanterix) were used to measure big tau and BD-tau in serum from patients with Guillain–Barré syndrome (GBS, n=81), Miller Fisher syndrome (MFS, n=20), Charcot–Marie–Tooth disease (CMT, n=102), chronic inflammatory demyelinating polyneuropathy (CIDP, n=43), MS (n=159), AD (n=20), and HC (n=41). NfL was measured in patients with neuropathies using an SR-X Single Molecule Array analyser (Quanterix).

**RESULTS:** Serum big tau levels were higher in GBS than in AD (11.4 vs 2.4 pg/mL, p<0.0001) and MS (11.4 vs 9.0 pg/mL, p=0.01), and similar to CIDP and CMT. Contrarily, serum BD-tau levels in GBS were higher than in CIDP (3.0 vs 2.3 pg/mL, p=0.006) and MS (3.0 vs 1.7 pg/mL, p<0.0001), but similar to CMT, and lower than in AD (3.0 vs 9.8 pg/mL, p<0.0001). Serum NfL levels were higher in GBS than in CIDP (32.5 vs 13.0 pg/mL, p=0.0002), CMT (32.5 vs 12.3 pg/mL, p<0.0001), and HC (32.5 vs 7.6 pg/mL, p<0.0001). Compared with GBS, MFS patients showed higher BD-tau (12.7 vs 3.0 pg/mL, p=0.003), lower big tau (5.4 vs 11.4 pg/mL, p=0.002), and higher NfL levels, although the latter did not reach statistical significance (118.3 vs 32.5 pg/mL, p=0.16). The NfL/big tau ratio was significantly higher in MFS than in GBS, CIDP, and CMT. In GBS, BD-tau correlated with early clinical severity (MRC at 1 week; I-RODS at 4 weeks; maximum GBS-DS and GBS-DS at 4 weeks), whereas neither tau biomarker showed long-term clinical correlations. Higher BD-tau and big tau levels were associated with the need for mechanical ventilation (BD-tau: 8.6 vs 2.9 pg/mL, p=0.019; big tau: 19.7 vs 10.7 pg/mL, p=0.007), while higher BD-tau levels were associated with mortality (10.9 vs 2.9 pg/mL, p=0.003).

**CONCLUSIONS:** Higher big tau levels in peripheral neuropathies than in CNS diseases support its role as a PNS-specific biomarker. In MFS, increased serum BD-tau, reduced big tau, and an elevated NfL/big tau ratio suggest CNS involvement with relative preservation of the PNS.

## INTRODUCTION

One of the most important unmet needs in peripheral nerve diseases is the identification of a specific biomarker for neuropathies that improves prognostic stratification, facilitates monitoring disease activity, and enables assessment of treatment response. Neuropathies are usually diagnosed based on clinical and electrophysiological criteria, but nerve conduction studies (NCS) are often poorly responsive to treatment, particularly in patients with severe axonal damage. Other available biomarkers, such as nerve imaging with magnetic resonance imaging (MRI) or ultrasound, lack specificity. Finding a reliable biomarker for peripheral nervous system (PNS) diseases is particularly challenging because peripheral nerve tissue constitutes only a small fraction of the total body volume, resulting in low concentrations of related proteins in the blood. This challenge is further compounded by the variable time course of nerve injury, which can be acute, as in Guillain-Barré Syndrome (GBS), or very prolonged over time, as in hereditary neuropathies^1^.

The development of ultrasensitive technologies capable of detecting biomarkers at very low concentrations in blood^2^ has enabled measurement of neurofilament light chain (NfL) levels across a large number of neurological diseases^3^, including several PNS disorders such as GBS^4,5^, chronic inflammatory demyelinating polyneuropathy (CIDP)^6–8^, inherited neuropathies^9,10^, and hereditary transthyretin-mediated amyloidosis with polyneuropathy^11,12^. Despite these advances, NfL is a general marker of neurodegeneration rather than a PNS-specific biomarker, and its levels can be influenced by age or comorbid central nervous system (CNS) involvement. In addition, its sensitivity remains uncertain in neuropathies with slow axonal damage, such as inherited neuropathies.

More recently, peripherin, an intermediate filament protein expressed in peripheral nerve axons, has been proposed as a promising biomarker for PNS axonal damage^13^. However, peripherin is also present in the pons and medulla oblongata, spinal cord, the cerebellum, and the corticospinal tract of the brainstem^14^, as well as in some peripheral organs, especially the gastrointestinal tract and lymph nodes, which may explain why its levels are not consistently higher in the PNS compared to certain CNS diseases^13^. Periaxin has also been described as a potential biomarker of peripheral nerve demyelination^13^, distinguishing peripheral from CNS diseases and identifying patients with active disease in CIDP, but its clinical value remains to be confirmed in other cohorts. This highlights the need to explore other molecules with stronger peripheral specificity.

A high-molecular-weight isoform of tau protein, known as big tau, is predominantly expressed in the PNS^15–17^. Tau is a microtubule-associated protein that stabilizes microtubules, regulates their dynamics, and influences axonal transport^18^. The *MAPT* gene, which encodes tau, is expressed as tissue- and organ-specific isoforms (45–65 kDa) through alternative splicing. Six major isoforms are primarily expressed in the brain. In contrast, the big tau isoform (∼110 kDa) contains a large exon (4a) and is the main tau variant in the adult PNS, as well as in neurons projecting to the periphery, such as dorsal root ganglia and ventral horn motoneurons ^15,19,20^. Recent studies have shown that big tau accounts for approximately 60% of total tau in the PNS, compared to only 1% in the brain, where it is primarily localized in the cerebellum^21^. Additionally, big tau is also expressed in non-neural tissues, such as the heart, kidney, and liver ^19,22^. Big tau appears to be relatively resistant to aggregation, yet it has received far less attention than the low-molecular-weight tau isoforms, which have been studied much more extensively in the context of tau biology and neurodegeneration. As a result, despite its potentially important distinct structural and functional properties, big tau remains comparatively poorly characterized, and its aggregation behavior, physiological roles, and pathological relevance are still not well understood^23^.

Recent methodological advances have enabled the ultrasensitive detection of tau isoforms. Among these, recently developed blood-based assays measure brain-derived tau (BD-tau)—all isoforms lacking exon 4a—which is increased in Alzheimer’s disease (AD). These assays improve the biomarker utility of plasma total tau while showing specificity for AD-type neurodegeneration^24^.

A major challenge in PNS diseases is the absence of a reliable, specific biomarker to improve monitoring of disease activity and treatment outcomes, and to serve as a suitable endpoint in early clinical trial development, which currently relies solely on clinical scoring. In this context, the present study aims to investigate the levels of big tau, BD-tau, and NfL in patients with neuropathies and to compare them with those in patients with CNS diseases.

## MATERIALS AND METHODS

### Study design and patients

We collected data from 81 GBS patients across 11 Spanish centers with samples at baseline, at one year in 25 patients, and at different time points in 13 patients. Patients fulfilling GBS diagnostic criteria were included within 2 weeks from disease onset. Patients were enrolled between February 2013 and January 2019. We also collected samples from 20 anti-GQ1b/GT1a positive Miller Fisher syndrome (MFS) patients referred to our hospital for antiganglioside antibody testing; 102 Charcot-Marie-Tooth (CMT) patients from 3 different Spanish centers, 43 CIDP patients from Hospital de la Santa Creu i Sant Pau, 159 MS patients from Hospital de la Santa Creu i Sant Pau, and 41 healthy controls (HC) from Hospital de la Santa Creu i Sant Pau. The AD group (*n* = 20) included serum samples from neurochemically defined Alzheimer’s disease individuals from the Sahlgrenska University Hospital, Sweden. Participants were selected based on their CSF biomarker profile (CSF Aβ42 < 530 pg/ml, CSF p-tau > 60 pg/ml and CSF t-tau > 350 pg/ml), and they had no evidence of other neurological conditions based on routine clinical and laboratory assessments. Demographic and clinical data for GBS were prospectively collected, and for MFS, CMT, CIDP, MS, and AD, retrospectively collected.

Serum samples were aliquoted and stored at −80°C until needed. All patients provided written informed consent to participate in the study. Written informed consent was obtained from all participants in accordance with the Declaration of Helsinki. Participation in the study was performed under a protocol approved by the Ethics Committee of the Hospital de la Santa Creu i Sant Pau (IIBSP-NAI2022-88). Sample collection from AD patients was approved by the ethics committees at the University of Gothenburg (#EPN140811). All experiments were conducted following the relevant guidelines and regulations.

### BD-tau and big tau measurements

Serum BD-tau was measured blinded on the Simoa HD-X using an in-house assay at the University of Gothenburg, Mölndal, Sweden, as previously described^24^. Serum Big tau was measured with a preliminary immunoassay design on the Simoa HD-X platform (**Fig. 1**). Signal variations within and between analytical runs were assessed using internal quality control samples analysed in duplicates at the beginning and the end of each run. The intra- and inter-assay coefficients of variation remained below 15% for both big tau and BD-tau based on internal quality controls.

**Figure 1.**
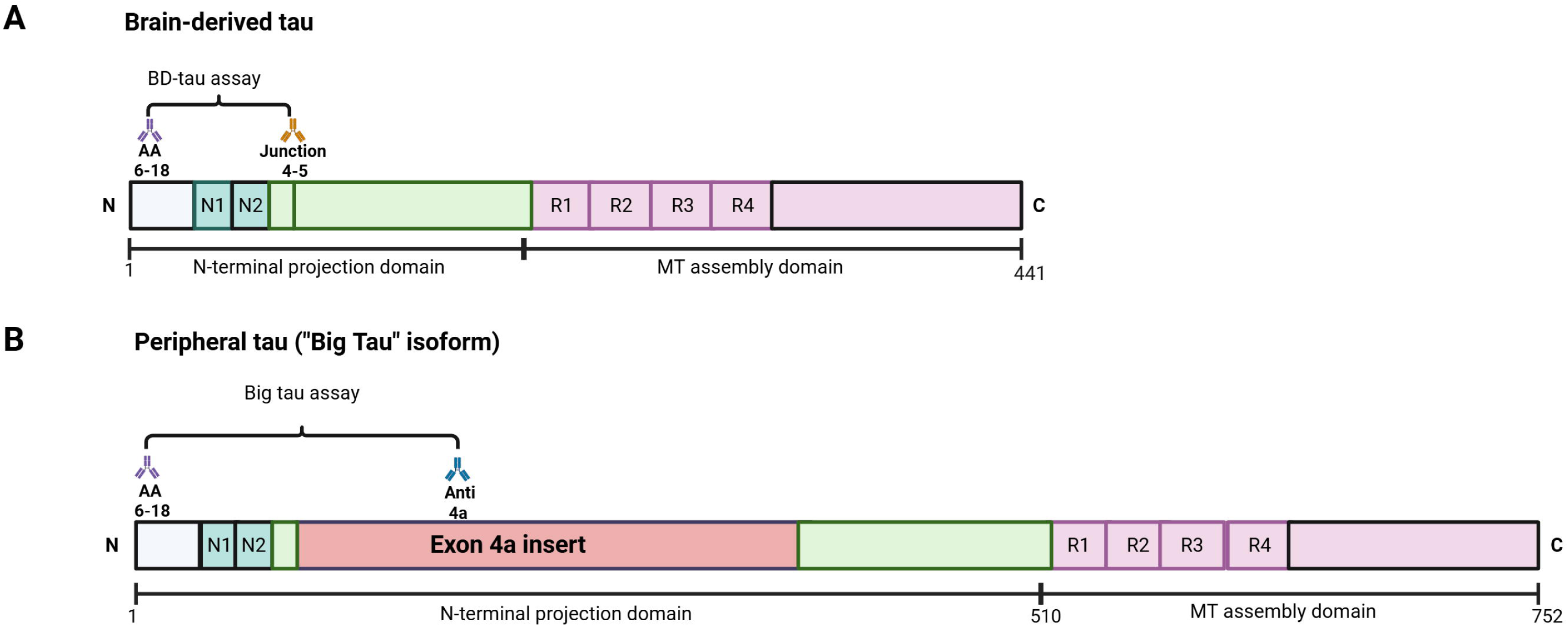
Schematic representation of the Brain-derived tau isoform (A, BD-tau) and the high molecular weight “Big tau” isoform (B). The BD-tau assay was specifically designed to target a contiguous peptide sequence at the junction of exons 4 and 5, a feature unique to the low-molecular weight isoforms of tau. In contrast, the big tau assay used an anti-exon 4a antibody raised against a sequence unique to the peripherally expressed big tau isoform. Adapted from Gonzalez-Ortiz et al. *Brain*, 2023. Created in https://BioRender.com.

### Simoa NfL measurements

Measurement of serum NfL levels was performed in peripheral nerve disorders using the Simoa Nf-light® kit on the SR-X immunoassay analyzer, Simoa™ (Quanterix Corp, Boston, MA, USA). Samples were analyzed in duplicates following the manufacturer’s instructions and standard procedures. Samples were analyzed at baseline and follow-up when available. All NfL values were within the linear ranges of the assays. The intra- and inter-assay coefficients of variation were 5.3% and 13.9%, respectively. NfL z-scores were calculated using the NfL Reference App^25^. Of note, NfL levels were not analyzed in the CNS disease groups (AD and MS), as the biomarker profile of NfL in these conditions is already robustly characterized and well-established in the literature.

### Statistical analysis

Continuous variables are described by mean ± standard deviation (SD) or median [interquartile range (IQR)]. Categorical variables are expressed as percentages. Univariate analysis was performed using the chi-squared test or Fisher’s exact test for dichotomous variables. Continuous variables were analyzed with the t-test or the Mann– Whitney U test when appropriate. The Kruskal-Wallis test was used to compare different groups. The Wilcoxon matched pairs signed rank test was used to compare biomarkers at baseline and at different time points. We used Spearman’s coefficient to assess the correlation between variables.

Statistical significance for all analyses was set at 0.05 (two-sided). The analysis was carried out in SPSS 32.0 (SPSS Inc., Chicago, Illinois, USA) and GraphPad Prism v8.

## RESULTS

### Demographic characteristics

We included patients with peripheral neuropathies (81 GBS, 20 MFS, 102 CMT and 43 CIDP), patients with CNS diseases (159 MS and 20 AD) and 41 HC. The mean age of patients was 47.9 years (SD 16.3), and 256 (58.2%) were female. Age and sex by group are shown in **Table 1**.

**Table 1.**
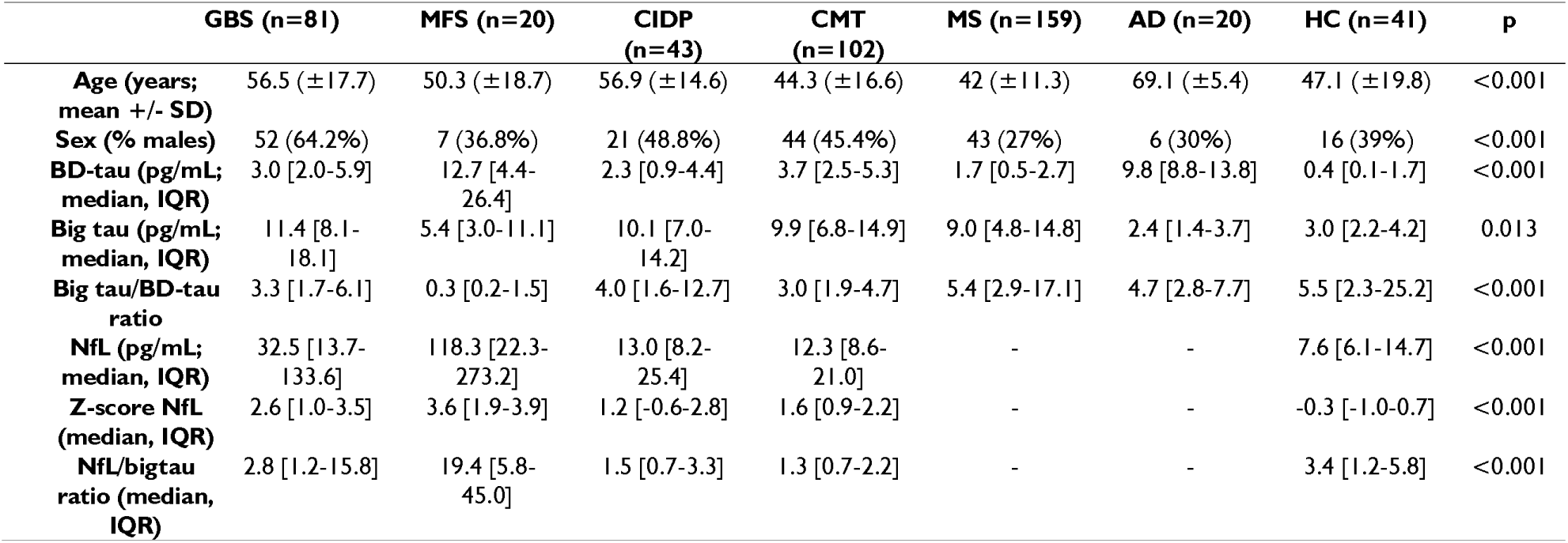
Biomarkers (BD-tau, big tau, big tau/BD-tau ratio, NfL and NfL/big tau ratio) in patients with GBS, CIDP, CMT, MS, AD and HC. AD: Alzheimer’s Disease; BD-tau: Brain-derived tau; CIDP: chronic inflammatory demyelinating polyneuropathy, CMT: Charcot-Marie-Tooth; GBS: Guillain-Barré syndrome; HC: Healthy controls, IQR: interquartile range; MS: Multiple Sclerosis; NfL: neurofilament light chain; SD: Standard deviation.

### BD-tau and big tau levels

BD-tau was significantly associated with age (r=0.13; p=0.009) across all patients; however, this correlation was not observed in the HC group (n=41; r=0.21, p=0.225). Big tau was not associated with age (r=-0.11, p=0.5). Neither biomarker was associated with gender.

GBS patients had significantly higher big tau levels than AD (11.4 vs 2.4 pg/mL, p<0.0001), MS patients (9.0 pg/mL, p=0.01) and HC (3.0 pg/mL, p<0.0001), and similar levels to CIDP or CMT patients (10.1 and 9.9 pg/mL, respectively) (**Fig. 2A**, **Table 1**). GBS patients had significantly higher BD-tau levels than CIDP (3.0 vs 2.3 pg/mL, p=0.03), MS patients (1.7 pg/mL, p<0.0001), and HC (0.4 pg/mL, p<0.0001) but similar levels to CMT patients (3.7 pg/mL) and lower levels than AD (9.8 pg/mL, p<0.0001) (**Fig. 2B**, **Table 1**). GBS patients had a lower big tau/BD-tau ratio than MS (3.3 vs 5.4, p>0.0001), AD (4.7, p=0.04) and HC (5.5, p=0.0007) and similar levels to CIDP and CMT patients (4.0 and 3.0, respectively) (**Fig. 2C**, **Table 1**).

**Figure 2.**
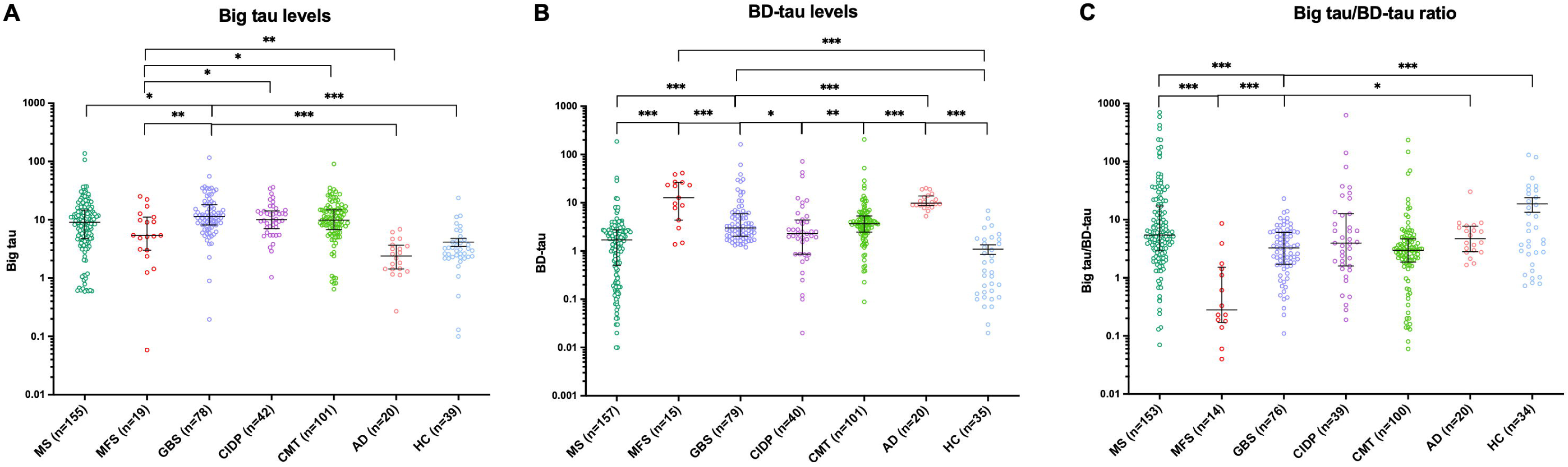
Big tau levels (A), BD-tau levels (B) and big tau/BD-tau ratio (Big tau/BD-tau) (C) in patients with GBS, CIDP, CMT, MS, AD and HC. The line in the centre represents the median value and the whiskers indicate the interquartile range. AD: Alzheimer’s Disease; BD-tau: Brain-derived tau; CIDP: chronic inflammatory demyelinating polyneuropathy, CMT: Charcot-Marie-Tooth; GBS: Guillain-Barré syndrome; HC: Healthy controls, MS: Multiple Sclerosis.

MFS patients had significantly lower big tau levels than GBS (5.4 vs 11.4 pg/mL, p=0.002), CIDP (10.1 pg/mL, p=0.01), CMT (9.9 pg/mL, p=0.026), higher levels than AD (2.4 pg/mL, p=0.003) and HC (3.0 pg/mL, p=0.016), and similar levels to MS (9.0 pg/mL, p=0.12) (**Fig. 2A**, **table 1**). MFS patients had significantly higher BD-tau levels than GBS (12.7 vs 3.0 pg/mL, p<0.0001), CMT (3.7 pg/mL, p=0.0004), CIDP (2.3 pg/mL, p=0.0005), MS (1.7 pg/mL, p<0.0001) and HC (0.4 pg/mL, p<0.0001) and similar levels to AD patients (9.8 pg/mL, p=0.49) (**Fig. 2B**, **table 1**). MFS patients had the lowest big tau/BD-tau ratio of all diseases (p<0.0001) (**Fig. 2C**, **table 1**).

### NfL levels

NfL levels were exclusively measured in patients with neuropathies, as their values in CNS diseases (MS and AD) have been extensively characterized in previous studies. GBS patients had significantly higher NfL levels than CIDP (32.5 vs 13.0 pg/mL; p=0.0002), CMT (12.3 pg/mL; p<0.0001), and HC (7.6 pg/mL; p<0.0001) (**Table 1**, **Fig. 3A**). NfL was significantly associated with age (r=0.758, p<0.01). These results remain at the same level of significance when age-corrected NfL Z-scores are evaluated (**Table 1**).

**Figure 3.**
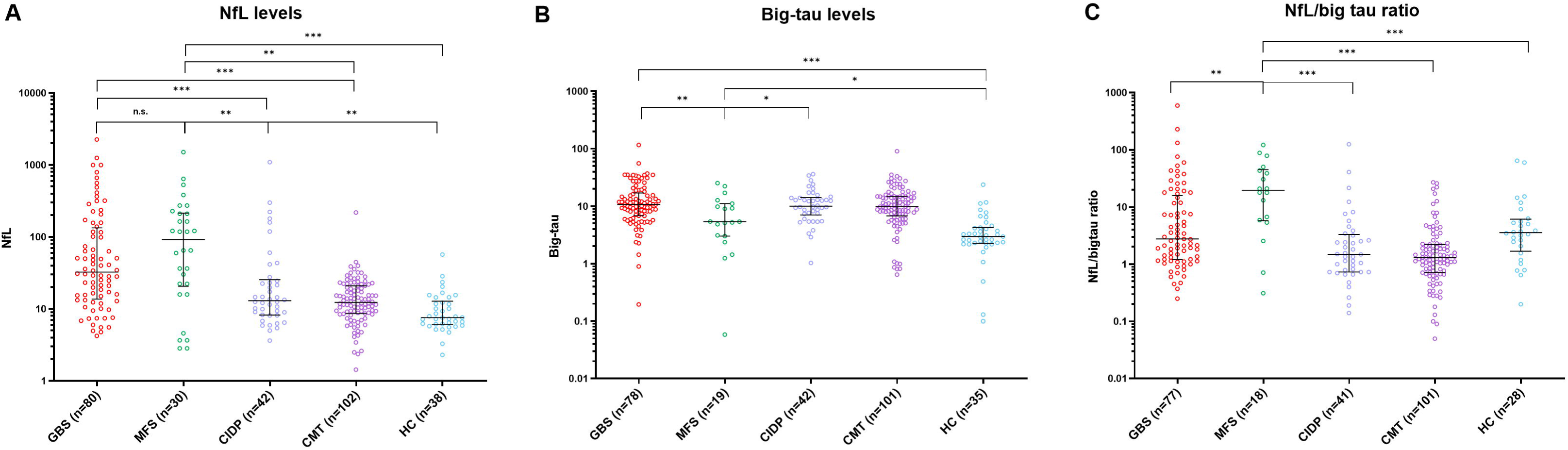
NfL (A), big tau (B) and NfL/big tau ratio (C) in patients with GBS, MFS, CIDP, CMT and HC. The line in the centre represents the median value and the whiskers indicate the interquartile range. CIDP: chronic inflammatory demyelinating polyneuropathy, CMT: Charcot-Marie-Tooth; GBS: Guillain-Barré syndrome; HC: Healthy controls, MFS: Miller Fisher Syndrome, NfL: neurofilament light chain.

MFS patients had higher median NfL values than GBS patients (118.3 vs 32.5 pg/mL, p=0.16), although the difference was not statistically significant. They also showed higher NfL levels than CIDP (13.0 pg/mL; p=0.002), CMT (12.3 pg/mL; p<0.001), and HC (7.6 pg/mL; p<0.0001). When analysing the ratio between NfL and big tau levels, MFS had a significantly higher NfL/big tau ratio than GBS (19.4 vs 2.8 pg/mL, p=0.004), CIDP (1.5 pg/mL, p<0.0001), and CMT patients (1.3 pg/mL, p<0.0001) (**Fig. 3C**, **Table 2**).

**Table 2.** Biomarkers in patients with MFS and GBS. BD-tau: Brain-derived tau; GBS: Guillain-Barré syndrome; IQR: interquartile range; MFS: Miller Fisher syndrome; NfL: neurofilament light chain; SD: Standard deviation.

|  | GBS (n=81) | MFS (n=20) | p |
| --- | --- | --- | --- |
| **Age (years; mean +/- SD)** | 56.5 (+/-17.7) | 50.3 (+/-18.7) | 0.20 |
| **Sex (% males)** | 52 (64.2%) | 7 (35%) | 0.03 |
| **BD-tau (pg/mL; median, IQR)** | 3.0 [2.0-5.9] | 12.7 [4.4-26.4] | 0.003 |
| **Big tau (pg/mL; median, IQR)** | 11.4 [8.1-18.1] | 5.4 [3.0-23.0] | 0.002 |
| **Big tau/BD-tau ratio** | 3.3 [1.7-6.1] | 0.3 [0.2-1.5] | <0.0001 |
| **NfL (pg/mL; median, IQR)** | 32.5 [13.7-133.6] | 118.3 [22.3-273.2] | 0.16 |
| **NfL percentile (median, IQR)** | 99.5 [76.8-99.9] | 99.9 [97.1-99.9] | 0.059 |
| **Z-scores (median, IQR)** | 2.6 [1.0-3.5] | 3.6 [1.9-4.0] | 0.15 |
| **NfL/bigtau ratio (median, IQR)** | 2.3 [1.2-15.8] | 19.4 [5.8-45.0] | 0.004 |

### Correlation analysis of NfL, BD-tau and big tau

Significant positive correlations were observed among the three biomarkers. Specifically, NfL levels showed a moderate association with BD-tau (r=0.338; p<0.001) and a weaker but significant correlation with big tau (r=0.229; p<0.001). Furthermore, big tau and BD-tau levels were positively correlated with each other (r=0.233; p<0.001); notably, this association was driven by the GBS (r=0.301, p<0.008) and MS patients (r=0.282, p<0.01) group, whereas no significant correlation was observed in the remaining groups.

### Association of biomarkers with disease characteristics

GBS patients with ganglioside antibodies had higher levels of all biomarkers (big tau 13.2 vs 8.3 pg/mL, p=0.001; BD-tau 3.8 vs 2.5 pg/mL, p=0.01; and NfL 49.3 vs 21.3, p=0.007; **Table 3**). Patients with the pure sensory variant had lower levels of big tau (5.8 vs 11.9 pg/mL, p=0.01) and BD-tau (1.7 vs 3.3 pg/mL, p=0.08) than those with the typical sensory-motor presentation. No differences were observed between patients with the pure motor variant and those with sensory-motor variants, nor were any significant differences found across NCS subtypes (**Table 3**).

**Table 3.** GBS characteristics, clinical scales and their correlation with biomarkers. BD-tau: Brain-derived tau; GBS: Guillain-Barré syndrome; GBS-DS: GBS Disability Scale; IQR: interquartile range; MRC Sum Score: Medical Research Council Sum Score; NCS: nerve conduction study; NfL: neurofilament light chain; SD: Standard deviation.

| GBS characteristics | Variables | BD-tau | Big tau | Big tau/BD-tau ratio | NfL |
| --- | --- | --- | --- | --- | --- |
| **Gangliosides** | n (%) | p=0.012 | p=0.001 | p=0.92 | p=0.007 |
| Yes (n, %) | 39 (49.4%) | 3.8 [2.4-7.6] | 13.2 [9.7-20.6] | 2.8 [1.5-6.2] | 49.3 [17.1-226.3] |
| No (n, %) | 38 (50.6%) | 2.5 [1.7-4.7] | 8.3 [5.7-12.3] | 3.3 [1.8-6.0] | 21.3 [8.4-81.1] |
| **GBS clinical variants** | n (%) | p=0.12 | p=0.054 | p=83 | p=0.015 |
| Sensori-motor | 61 (75.3%) | 3.3 [2.0-6.8] | 11.9 [8.5-18.2] | 2.6 [1.6-6.3] | 29.1 [13.7-63.3] |
| Pure sensory | 4 (5%) | 1.7 [1.6-2.3] | 5.8 [2.1-8.9] | 3.4 [1.1-5.4] | 7.7 [4.4-1704.6] |
| Pure motor | 16 (19.7%) | 2.4 [2.2-4.4] | 10.2 [8.2-19.3] | 3.7 [2.3-5.4] | 47.8 [28.0-523.7] |
| **NCS classification** | n (%) | p=0.34 | p=0.29 | p=0.44 | p=0.036 |
| AIDP | 56 (69.1%) | 2.9 [1.9-6.1] | 11.9 [8.2-19.4] | 3.5 [1.9-6.5] | 25.2 [12.9-63.4] |
| AMAN | 10 (12.3%) | 3.6 [2.1-5.8] | 10.8 [8.5-20.1] | 3.7 [2-4.3] | 253.8 [90.4-746.6] |
| AMSAN | 5 (6.2%) | 3.3 [2.9-12.6] | 13.2 [4.8-15.4] | 1.2 [0.7-4.1] | 101.2 [19.4-237.2] |
| Equivocal | 7 (8.6%) | 4.7 [2.6-5.8] | 8.6 [5.2-12.3] | 1.8 [0.9-6.1] | 36.1 [28.2-63.1] |
| Normal | 2 (2.5%) | 1.8 | 6.2 | 3.4 | 17 |
| **Clinical scales** | Median [IQR] |  |  |  |  |
| Baseline MRC Sum Score | 50 [42-56] | r=-0.27; p=0.015 | r=-0.17; p=0.14 | r=0.07; p=0.55 | r=-0.46; p<0.001 |
| MRC Sum Score at 1 week | 51 [42-58] | r=-0.35; p=0.002 | r=-0.18; p=0.12 | r=0.17; p=0.16 | r=-0.38; p<0.001 |
| I-RODS at 4 weeks | 34 [14-45] | r=-0.35; p=0.004 | r=-0.20; p=0.12 | r=0.19; p=0.15 | r=-0.45; p<0.001 |
| I-RODS at 6 months | 44 [33-48] | r=-0.19; p=0.14 | r=-0.20; p=0.12 | r=0.07; p=0.24 | r=-0.43; p<0.001 |
| I-RODS at 1 year | 45 [35-48] | r=-0.20; p=0.11 | r=-0.043; p=0.74 | r=0.15; p=0.24 | r=-0.43; p<0.001 |
| Final I-RODS | 46 [37-48] | r=0.21; p=0.089 | r=-0.11; p=0.39 | r=0.11; p=0.4 | r=-0.40; p<0.001 |
| Initial GBS-DS | 3 [2-4] | r=0.21; p=0.069 | r=0.13; p=0.26 | r=-0.12; p=0.31 | r=0.38; p<0.001 |
| Maximum GBS-DS | 4 [2-4] | r=0.30; p=0.008 | r=0.15; p=0.18 | r=-0.18; p=0.12 | r=0.32; p=0.004 |
| GBS-DS at 4 weeks | 2 [1-4] | r=0.34; p=0.002 | r=0.19; p=0.1 | r=-0.17; p=0.14 | r=0.34; p=0.002 |
| **Outcomes** | n (%) |  |  |  |  |
| Ability to walk at 6 months |  | p=0.30 | p=0.87 | p=0.47 | p=0.025 |
| Yes | 63 (87.5%) | 2.8 [1.8-5.8] | 11.1 [8.2-18.4] | 3.5 [1.8-6.1] | 26.7 [12.8-68.4] |
| No | 9 (12.5%) | 3.2 [2.5-16.9] | 11.5 [8.0-18.4] | 2.3 [1.2-6.3] | 156.7 [35.4-641.0] |
| Ability to run at 1 year |  | p=0.54 | p=0.92 | p=0.54 | p<0.001 |
| Yes | 47 (66.2%) | 2.9 [1.7-5.6] | 11.6 [6.8-18.4] | 3.5 [1.7-6.3] | 22.3 [10.3-51.6] |
| No | 24 (33.8%) | 3.2 [2.3-6.4] | 10.0 [8.7-13.4] | 3.1 [1.8-5.4] | 64.8 [35.0-400.3] |
| Mechanical ventilation |  | p=0.019 | p=0.007 | p=0.4 | p=0.4 |
| Yes | 8 (9.9%) | 8.6 [2.9-30.4] | 19.7 [13.5-31.5] | 2.2 [0.7-6.8] | 45.0 [14.7-447.2] |
| No | 73 (90.1%) | 2.9 [1.9-5.1] | 10.7 [7.7-15.4] | 3.5 [1.8-6.1] | 29.2 [13.3-117.5] |
| Death |  | p=0.003 | p=0.69 | p=0.11 | p=0.013 |
| Yes | 5 (6.2%) | 10.9 [6.1-29.1] | 11.9 [8.4-22.6] | 0.90 [0.5-2.2] | 50.0 [41.6-404.7] |
| No | 76 (93.8%) | 2.9 [2.0-5.1] | 10.8 [8.1-17.6] | 3.5 [1.8-6.1] | 29.2 [13.2-117.5] |

BD-tau correlated with clinical scales at disease onset in GBS patients: for MRC Sum Score at 1 week (r=-0.35, p=0.002) and for I-RODS at 4 weeks (r=-0.35, p=0.004) for maximum GBS-DS (r=0.30, p=0.008) and GBS-DS at 4 weeks (r=0.34, p=0.002). Patients that needed ventilation had higher BD-tau and big tau levels (8.6 vs 2.9 pg/mL, p=0.019 and 19.7 vs 10.7 pg/mL, p=0.007, respectively) and patients who deceased had higher BD-tau levels (10.9 vs 2.9 pg/mL; p=0.003). Big tau showed only a trend to be associated with clinical scales at the disease onset (**Table 3**). Unlike NfL levels, neither tau biomarker showed long-term clinical correlations, except for a trend between baseline BD-tau and the last-visit I-RODs score (r=0.21; p=0.089) (**Table 3**).

No differences were found between axonal and demyelinating variants in CMT patients regarding BD-tau, big tau, NfL levels, big tau/BD-tau ratio or NfL/big tau ratio. In patients with MS, we observed a trend towards a correlation between EDSS and BD-tau (r=0.15, p=0.07). Naïve patients had higher BD-tau and big tau levels than those receiving disease-modifying treatments (2.7 vs 1.5 pg/mL, p=0.044, and 11.1 vs 8.4 pg/mL, p=0.038, respectively). No differences were found among MS subtypes, disease duration, or the presence of spinal cord lesions across biomarkers (**Supplementary Table 1**).

### Longitudinal serum biomarker dynamics

BD-tau and big tau levels decreased at 1 year in GBS patients with higher baseline levels but remained stable or increased slightly at follow-up in most GBS patients (**Fig. 4**). Big tau levels increased at weeks 1 and 2 to subsequently decrease up to 1 year (**Fig. 5**).

**Figure 4.**
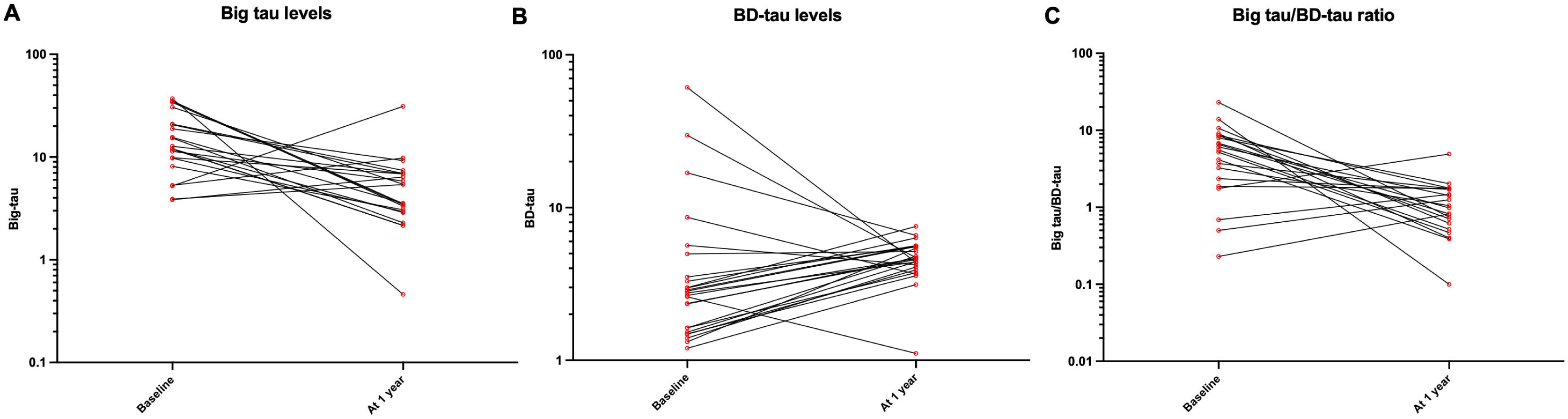
Big tau (A), BD-tau levels (B) and big tau/BD-tau ratio (C) in GBS patients at 1 year (n=25). BD-tau: Brain-derived tau, GBS: Guillain-Barré Syndrome.

**Figure 5.**
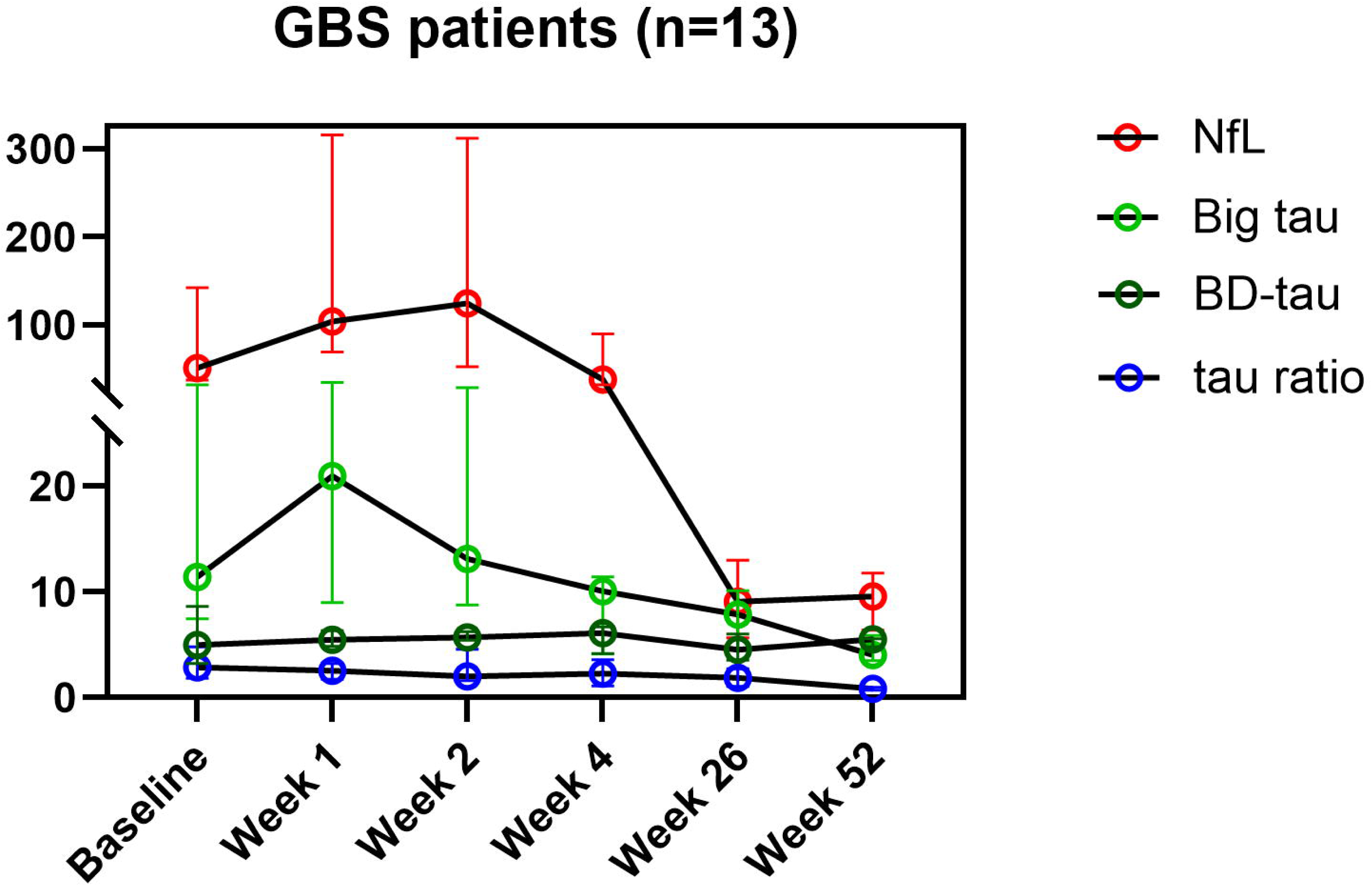
Big tau, BD-tau, big tau/BD-tau ratio and NfL kinetics in GBS patients (n=13). BD-tau: Brain-derived tau, GBS: Guillain-Barré Syndrome, NfL: neurofilament light chain.

## DISCUSSION

Our results show that patients with neuropathies have higher levels of big tau than those with CNS diseases, with GBS patients presenting higher BD-tau levels than those with CIDP or MS. Furthermore, MFS displayed a distinct biomarker profile —higher BD-tau, lower big tau, and a higher NfL/big tau ratio— suggesting CNS involvement and relative preservation of the PNS in those patients. Finally, BD-tau correlated with clinical scales early in the disease in GBS patients, and normalized in patients with GBS upon disease resolution and in MS patients after treatment initiation.

To our knowledge, this is the first study measuring big tau levels in patients with different peripheral and central neurological diseases. Previous studies have focused on total tau or other axonal biomarkers, such as neurofilament heavy chain (NfH), particularly in the CSF of GBS patients, and have shown that higher CSF tau and NfH levels are associated with a worse prognosis (inability to walk independently)^25,26^. One study found correlations between CSF tau and NfH and the GBS Disability Scale (GBS-DS) in patients with acute motor axonal neuropathy (AMAN), compared with other neurological pathologies, and a correlation between CSF S100B and GBS-DS in patients with acute inflammatory demyelinating polyradiculoneuropathy (AIDP)^27^. More recently, NfL and total-tau were evaluated in CSF and plasma from patients with GBS, CIDP, paraproteinemia-related demyelinating polyneuropathy and multifocal motor neuropathy (MMN), showing that both acute and chronic inflammatory neuropathies are associated with higher levels of NfL and total tau in CSF and plasma; however, only NfL was validated as a prognostic biomarker in GBS^26–28^.

Despite being identified long ago, big tau remains poorly understood, leaving an important gap in knowledge about its potential role as a biomarker for PNS disorders^19,29^. Although it is the predominant tau isoform in the PNS, it is also expressed in selective regions of the CNS, including the optic nerve, cerebellum and dorsal root ganglia^15,16^. This distribution might explain why MS patients have higher big tau levels than AD patients in our study.

Surprisingly, we found that GBS patients have higher BD-tau levels than those in some other CNS diseases, including MS. There is only one article describing high BD-tau levels in GBS^32^, but it does not compare them with those in different neurological diseases. Unlike our findings, they observed a correlation between long-term outcomes (ONLS at 1 year; GBS-DS> 2 at 1 year) and BD-tau levels, suggesting that CNS involvement influences recovery. Although GBS is primarily a PNS disorder, CNS involvement may occur to varying degrees, as inflammatory changes in GBS can reach the intrathecal segments of nerve roots and may even extend proximally to involve spinal anterior horn cells^30–32^. Some reports of GBS cases presenting with hyperreflexia point to potential involvement of spinal interneurons or the corticospinal tract, especially in patients with AMAN^33,34^. This could explain the elevated BD-tau levels in some of our GBS patients (although no significant increase in BD-tau was found in AMAN or pure motor variants in our cohort, which might be explained by the limited sample size in these subgroups). Furthermore, we found that elevated BD-tau levels were significantly associated with the need for mechanical ventilation and mortality, which raises another critical consideration. It is plausible that severe systemic disease requiring intensive care unit admission—along with its associated complications, such as critical illness-related brain dysfunction or transient hypoxia—could induce secondary CNS injury, thereby confounding or artificially elevating BD-tau levels independently of the primary GBS pathophysiology. The higher big tau levels observed in mechanically ventilated patients confirm the more severe nature of GBS in those cases.

Additionally, CNS involvement is particularly relevant in clinical variants such as Bickerstaff brainstem encephalitis (BBE) and MFS, both associated with GQ1b antibodies^35–37^. In this context, the biomarker profile we observed in MFS patients appears particularly informative. There has been controversy regarding the presence of CNS involvement in MFS patients^38^. While in BBE decreased consciousness and hyperreflexia make CNS involvement more plausible, in MFS, the origin of the ataxia and ophthalmoparesis has been less clear. Early descriptions proposed cerebellar dysfunction as the cause for ataxia and brainstem lesions for areflexia^39^, but the identification of GQ1b antibodies in the paranodal regions of ocular motor nerves^40^ shifted attention towards PNS involvement. Nonetheless, GQ1b epitopes have also been identified in the cerebellum^41^. Overlapping clinical features and paraclinical findings between BBE and MFS further support this possibility^35^. Additionally, although brain MRI and EEG abnormalities are more frequent in BBE, similar changes were found in a subset of MFS patients, despite preserved consciousness, supporting occasional CNS involvement^35^. One electrophysiological study also suggested cerebellar fibre system involvement in MFS^41^, and PET findings with glucose hypermetabolism in the cerebellum and brainstem in patients with MFS have been reported^42^. Our own previous work showed higher NfL levels in MFS patients in comparison with other GBS variants, despite normal peripheral nerve electrophysiology^28^, suggesting preganglionic root involvement or cerebellar damage. Taken together, the elevated NfL, higher BD-tau, and lower big tau observed in our study strengthen the argument for CNS involvement in MFS.

Another unexpected finding was that the damage reflected by big tau in patients with GBS was similar to that in CMT or CIDP. This observation may reflect a ceiling effect in the big tau assay, although other potential factors may explain this phenomenon. For example, circulating big tau may also originate from other tissues, since its expression has been reported in the myocardium, liver, and kidney^19,22,43^and part of the circulating levels may not be PNS-derived. Potential confounders, such as kidney failure—which is relatively common in elderly patients—should also be considered. Furthermore, our longitudinal data show that big tau peaks at week 1 after disease onset in GBS (similar to what happens with sNfL^44^), indicating that big tau is nerve-specific and that this timepoint may be optimal for exploring correlations with clinical outcomes, comparisons with other biomarkers and level comparisons across different diseases.

This study has several limitations. First, its retrospective design inherently limits the ability to control for potential confounding variables and to draw causal inferences. Second, comorbidities and other relevant clinical factors were not systematically considered in big tau and BD-tau. Third, temporal sampling was incomplete, and dynamic changes over time could not be systematically assessed for all patients. Finally, although big tau and BD-tau appear to be new axonal biomarkers for distinguishing between CNS and PNS disorders, larger studies should be conducted to clarify the temporal dynamics and to fully elucidate the value of big tau in neuropathies.

In conclusion, our study is the first to assess big tau as a potential biomarker for PNS disorders. We demonstrate that neuropathies have higher big tau levels than CNS diseases. Although long-term prognostic value was not observed, a distinct biomarker profile in MFS suggests central involvement in this condition, highlighting the potential of combined biomarker strategies to improve diagnostic accuracy in inflammatory neuropathies.

## Data Availability

Data are available upon reasonable request. All data relevant to the study are included in the article or uploaded as supplementary information. Anonymized data not published within this article will be made available by request from any qualified investigator.

## Acknowledgements

Some authors of this publication are members of the European Reference Network for rare neuromuscular diseases (EURO-NMD).

## ABBREVIATIONS

AD: Alzheimer’s diseas
BD-tau: Brain-derived tau
CIDP: Chronic Inflammatory Demyelinating Polyneuropathy
CMT: Charcot-Marie-Tooth
GBS: Guillain-Barré Syndrome
HC: healthy controls
I-RODS: Inflammatory Raschbuilt Overall Disability Scale
IQR: interquartile range
MFS: Miller Fisher syndrome
MRC Sum Score: Medical Research Council Sum Score
MS: multiple sclerosis
MT: microtubule
NCS: nerve conduction studies
NfL: neurofilament light chain
PNS: peripheral nerve system
SD: Standard deviation.

---

**Supplementary table 1.** Clinical characteristics in patients with MS and their correlation with biomarkers. AAR: annualized relapse rate; EDSS: Expanded Disability Status Scale, IQR: interquartilic range; MS: Multiple Sclerosis

| Clinical characteristics MS (n=159) |  | BD-tau | Big tau | Big tau/BD-tau ratio |
| --- | --- | --- | --- | --- |
| Age | 43.3 (+/-11.3) | r=0.007, p=0.93 | r=-0.27; p=0.74 | r=-0.04; 0.63 |
| MS subtype |  | p=0.83 | p=0.69 | p=0.56 |
| RRMS | 128 (80.5%) | 1.6 [0.4-2.7] | 9.0 [4.7-14.7] | 5.7 [2.9-17.0] |
| SPMS | 13 (8.2%) | 1.8 [0.3-3.5] | 9.5 [6.8-21.6] | 8.0 [3.0-39.1] |
| PPMS | 18 (11.3%) | 1.9 [1.2-2.9] | 8.6 [6.4-14.5] | 5.1 [2.9-8.8] |
| Sex |  | p=0.19 | p=0.88 | p=0.12 |
| Male (n, %) | 43 (27%) | 1.3 [0.2-2.7] | 8.7 [4.2-14.1] | 6.7 [3.8-31.4] |
| Female (n, %) | 116 (73%) | 1.9 [0.6-2.8] | 9.1 [4.9-16.2] | 5.2 [2.7-11.6] |
| EDSS (median, IQR) | 2 [1-4] | r=0.15, p=0.07 | r=0.036, p=0.66 | r=-0.12, p=0.15 |
| Relapse last year (n, %) |  | p=0.68 | p=0.74 | p=0.44 |
| Yes | 43 (27%) | 1.7 [0.6-3.0] | 8.7 [4.3-14.1] | 4.7 [3.2-7.9] |
| No | 116 (73%) | 1.7 [0.5-2.7] | 9.1 [4.9-15.3] | 6.0 [2.7-19.1] |
| ARR (median, IQR) | 0.25 [0-0.5] | r=0.001, p=0.99 | r=-0.018; p=0.84 | r=0.005, p=0.96 |
| Disease duration (median, IQR) | 5.5 [2-11] | r=0.010, p=0.90 | r=-0.031, p=0.71 | r=-0.080, p=0.33 |
| Available spinal cord MRI |  | p=0.68 | p=0.53 | p=0.61 |
| Yes | 87 (54.7%) | 1.8 [0.5-2.7] | 9.6 [5.3-14.7] | 5.1 [3.0-16.1] |
| No | 72 (45.3%) | 1.6 [0.5-2.7] | 8.7 [4.3-16.0] | 6.4 [2.6-20.2] |
| Presence of spinal cord lesions (n=87) |  | 0.22 | 0.043 | 0.94 |
| Yes | 72 (82.8%) | 1.6 [0.4-2.7] | 8.6 [5.0-13.4] | 5.1 [3.0-17.6] |
| No | 15 (17.2%) | 2.2 [1.4-3.5] | 14.7 [11.2-26.6] | 4.9 [3.3-11.0] |
| Naïve (n, %) |  | p=0.044 | p=0.038 | p=0.91 |
| Yes | 30 (18.9%) | 2.7 [0.8-3.2] | 11.1 [8.5-16.2] | 5.1 [3.7-9.4] |
| No | 129 (81.1%) | 1.5 [0.5-2.6] | 8.4 [4.2-14.5] | 5.9 [2.7-17.8] |

